# Performance of antigen lateral flow devices in the United Kingdom during the Alpha, Delta, and Omicron waves of the SARS-CoV-2 pandemic

**DOI:** 10.1101/2022.11.29.22282899

**Authors:** David Eyre, Matthias Futschik, Sarah Tunkel, Jia Wei, Joanna Cole-Hamilton, Rida Saquib, Nick Germanacos, Andrew R Dodgson, Paul E Klapper, Malur Sudhanva, Chris Kenny, Peter Marks, Edward Blandford, Susan Hopkins, Tim Peto, Tom Fowler

**Affiliations:** Big Data Institute, Nuffield Department of Population Health, University of Oxford, UK; NIHR Oxford Biomedical Research Centre, University of Oxford, UK; NIHR Health Protection Research Unit in Healthcare Associated Infections and Antimicrobial Resistance, University of Oxford, UK; Public Health and Clinical Oversight (PHCO), UK Health Security Agency, London, UK; Nuffield Department of Medicine, University of Oxford, UK; UK Health Security Agency, Department of Microbiology, Manchester Public Health Laboratory, Manchester, UK; Division of Evolution, Infections and Genomics, University of Manchester, UK; South London Specialist Virology Centre, Infection Sciences, King’s College NHS Foundation Trust, London, UK; UK Health Security Agency, London, UK

## Abstract

**Background:** Antigen lateral flow devices (LFDs) have been widely used to control SARS-CoV-2. Changes in LFD sensitivity and detection of infectious individuals during the pandemic with successive variants, vaccination, and changes in LFD use are incompletely understood.

**Methods:** Paired LFD and PCR tests were collected from asymptomatic and symptomatic participants, across multiple settings in the UK between 04-November-2020 and 21-March-2022. Multivariable logistic regression was used to analyse LFD sensitivity and specificity, adjusting for viral load, LFD manufacturer, setting, age, sex, assistance, symptoms, vaccination, and variant. National contact tracing data were used to estimate the proportion of transmitting index cases (with ≥1 PCR/LFD-positive contact) potentially detectable by LFDs over time, accounting for viral load, variant, and symptom status.

**Findings:** 4131/75,382 (5.5%) participants were PCR-positive. Sensitivity vs. PCR was 63.2% (95%CI 61.7-64.6%) and specificity 99.71% (99.66-99.74%). Increased viral load was independently associated with being LFD-positive. There was no evidence LFD sensitivity differed between Delta vs. Alpha/pre-Alpha infections, but Omicron infections were more likely to be LFD positive. Sensitivity was higher in symptomatic participants, 68.7% (66.9-70.4%) than in asymptomatic participants, 52.8% (50.1-55.4%). 79.4% (68.6-81.3%) of index cases resulting in probable onward transmission with were estimated to have been detectable using LFDs, this proportion was relatively stable over time/variants, but lower in asymptomatic vs. symptomatic cases.

**Interpretation:** LFDs remained able to detect most SARS-CoV-2 infections throughout vaccine roll-out and different variants. LFDs can potentially detect most infections that transmit to others and reduce risks. However, performance is lower in asymptomatic compared to symptomatic individuals.

**Funding:** UK Government.

**Research in context:** *Evidence before this study:* Lateral flow devices (LFDs; i.e. rapid antigen detection devices) have been widely used for SARS-CoV-2 testing. However, due to their imperfect sensitivity when compared to PCR and a lack of a widely available gold standard proxy for infectiousness, the performance and use of LFDs has been a source of debate. We conducted a literature review in PubMed and bioRxiv/medRxiv for all studies examining the performance of lateral flow devices between 01 January 2020 and 31 October 2022. We used the search terms ‘SARS-CoV-2’/’COVID-19’ and ‘antigen’/’lateral flow test’/’lateral flow device’. Multiple studies have examined the sensitivity and specificity of LFDs, including several systematic reviews. However, the majority of the studies are based on pre-Alpha infections. Large studies examining the test accuracy for different variants, including Delta and Omicron, and following vaccination are limited.

*Added value of this study:* In this large national LFD evaluation programme, we compared the performance of three different LFDs relative to PCR in various settings. Compared to PCR testing, sensitivity was 63.2% (95%CI 61.7-64.6%) overall, and 71.6% (95%CI 69.8-73.4%) in unselected communitybased testing. Specificity was 99.71% (99.66-99.74%). LFDs were more likely to be positive as viral loads increased. LFD sensitivity was similar during Alpha/pre-Alpha and Delta periods but increased during the Omicron period. There was no association between sensitivity and vaccination status. Sensitivity was higher in symptomatic participants, 68.7% (66.9-70.4%) than in asymptomatic participants, 52.8% (50.1-55.4%). Using national contact tracing data, we estimated that 79.4% (68.6-81.3%) of index cases resulting in probable onward transmission (i.e. with ≥1 PCR/LFD-positive contact) were detectable using LFDs. Symptomatic index cases were more likely to be detected than asymptomatic index cases due to higher viral loads and better LFD performance at a given viral load. The proportion of index cases detected remained relatively stable over time and with successive variants, with a slight increase in the proportion of asymptomatic index cases detected during Omicron.

*Implications of all the available evidence:* Our data show that LFDs detect most SARS-CoV-2 infections, with findings broadly similar to those summarised in previous meta-analyses. We show that LFD performance has been relatively consistent throughout different variant-dominant phases of the pandemic and following the roll-out of vaccination. LFDs can detect most infections that transmit to others and can therefore be used as part of a risk reduction strategy. However, performance is lower in asymptomatic compared to symptomatic individuals and this needs to be considered when designing testing programmes.

## Introduction

Early detection of symptomatic and asymptomatic SARS-CoV-2 infections has been a key control measure during the COVID-19 pandemic. Rapid point-of-care antigen detection lateral flow devices (LFDs) have been widely in testing in the United Kingdom (UK),[1,2] including, at different times, for population-wide asymptomatic screening, and screening in specific groups such as healthcare workers, school-age children, and populations with increased incidence. LFDs have also been used to allow contacts of infected cases to continue to work or attend education, as well as prior to travel, attending events, and visiting residential care facilities.

However, LFDs have generated considerable scientific and policy debate.[3] They have imperfect sensitivity relative to PCR testing: ranging from <50% to >80%.[4–6] Sensitivity is greater in symptomatic vs. asymptomatic infection, e.g. 72% vs. 58%.[4] This had led to concerns that LFDs may miss some infections and paradoxically increase transmission if individuals testing falsely-negative reduce transmission precautions.[7] Conversely, LFDs need not detect and prevent all transmission to still have an effect at a population level; particularly where reproduction numbers are marginally over 1, an imperfect intervention may still be sufficient to control an outbreak.[8–11]

An additional concern is the lack of a widely-available proxy for an individual being infectious, as PCR-positivity may persist for days to weeks following the end of symptoms and/or infectiousness. It is therefore difficult to directly assess the sensitivity of LFDs in those who are infectious, which would be a better measure of their performance as a control intervention than sensitivity relative to PCR testing.[12] The likelihood of onward transmission is also related to index case viral load, with lower measured PCR cycle threshold (Ct) values (a proxy for higher viral loads in the tested individual) being associated with more secondary cases.[13,14]

LFDs are more sensitive as viral load increases,[4,14] such that performance may be better understood by a curve reporting sensitivity at various viral loads, rather than simply a single figure that also depends on the distribution of viral loads in the population studied. By combining index case PCR viral loads, contact tracing data, and LFD-viral load performance curves we have previously shown that LFDs would be expected to have detected most infections leading to onward transmission in the Alpha period in the UK.[14]

Here we present data on the performance of LFDs relative to PCR across multiple evaluations conducted in a range of settings during phases of the pandemic where the Alpha, Delta and Omicron variants dominated, to evaluate if performance has remained stable over time, including with any changes in population-wide proficiency and use of the tests, following vaccinations and with viral evolution. We combine our findings with contact tracing data to also evaluate the proportion of infectious cases detected by LFDs over time.

## Methods

### LFD evaluations: participants and samples

Prospective collection of paired LFD results and samples for PCR testing was undertaken to evaluate the performance of LFDs and their deployment, and for service quality assurance by NHS Test & Trace (now part of the UK Health Security Agency, UKHSA). Participants were asked to take a second test as well as their standard test specifically for the purposes of evaluating the LFDs and the testing programme. Prior to wider national deployment, the performance of Innova LFDs was evaluated in two field studies, referred to as “predeployment testing”. We analysed data collected from the start of the evaluation programme until the end of provision of free testing for the general public in March-2022.

Paired PCR and LFD testing was undertaken in several settings. Pre-deployment data were obtained from symptomatic testing sites, with a mixture of assisted and self-taken swabs. Subsequent settings included testing offered to asymptomatic individuals in the community (from the general public via testing sites and home testing; city-wide testing in Liverpool during a period of increased incidence[15]); targeted community testing (disproportionately impacted and under-served groups); schools (predominantly secondary); private and public sector workplaces; universities; and healthcare staff. Paired testing was also undertaken at local and regional symptomatic testing sites.

Participants were provided with instructions on how to perform testing. Two separate combined throat and/or nose swabs were obtained for testing (see Supplement). Swabs were self-taken. In assisted testing the LFD result was interpreted by a trained individual. In self-testing, the user (or relevant person on behalf of the user such as a parent/guardian/carer) self-swabbed and interpreted the test themselves without assistance. Evaluations were carried out in live testing services, therefore standard testing was always prioritised over the supplementary test, meaning there was no randomisation of swabbing order during sample collection. If standard testing was LFD, participants were asked to put the LFD to one side to develop, while providing the PCR sample (so that they were unaware of the result of either test at sampling). Participants or operatives (where testing was assisted) were asked to interpret LFD results as positive, negative, or void. Participants were unaware of PCR results at the time of LFD testing. Similarly, laboratories undertaking PCR testing were unaware of LFD results.

### LFD evaluation: sample assays

LFDs evaluated were the (i) Innova SARS-CoV-2 Lateral Flow Antigen Test (Innova in original packaging or repacked with individual buffer containers, also known as Biotime); (ii) Orient Gene COVID-19 Ag Rapid Test Cassette LFD antigen tests (Orient Gene); and (iii) Acon Flowflex SARS-CoV-2 antigen rapid test (Self-Testing) kit (Acon). PCR testing was undertaken by routine laboratories within the NHS Test and Trace laboratory network, and was performed predominantly using the Thermo Fisher SARS-CoV-2 TaqPath assay.

### Contact tracing data

National contact tracing data from England was obtained from NHS Test & Trace, as described previously.[13,14] Index cases were notified to the service following a positive community or healthcare-based SARS-CoV-2 PCR or LFD result. All index cases with a diagnostic PCR test performed using the Thermo Fisher SARS-CoV-2 TaqPath assay at three national testing “Lighthouse” laboratories in Milton Keynes, Alderley Park and Glasgow were eligible for inclusion.

Contacts (persons living in the same household, or in face-to-face distance from an index patient, within <1 m for ≥1 minute or within <2 m for ≥15 minutes) were eligible for inclusion in the study, provided they were only named by a single index patient in the 14 days either side of the index patient’s positive test. Contacts with any positive PCR or LFD result in the 1 to 10 days after the index patient’s positive test were considered to represent plausible transmission events. The 1 to 10 day period was chosen to enrich for contacts for whom the index patient was the most likely source of any infection, as previously described.[13,14]

### Statistical analysis

For LFD performance evaluations the infecting variant in PCR-positive infections was assigned based on sequencing/genotyping where available and if not, based on the dominant circulating variant (see Supplement). Real-time quantitative PCR cycle threshold (Ct) values were used to estimate SARS-CoV-2 viral loads in swab fluids in copies/mL using calibrant samples (Table S1).

PCR-positive samples were used for analyses of LFD sensitivity. Univariable and multivariable logistic regression was used to model the relationship between LFD positivity and log10 viral load and other covariates. Covariates included LFD device, study setting, assisted vs. self-testing, self-reported symptom status (symptomatic, i.e. fever, cough or anosmia/ageusia, otherwise asymptomatic), vaccination status by number of doses (0, 1, 2 or more doses) and viral variant (Alpha [B.1.1.7] / pre-Alpha [B.1.177], Delta [B.1.617.2], Omicron [BA.1 and BA.2]; or Other / Unknown). PCR-negative samples were used to analyse LFD specificity using univariable and multivariable logistic regression and the same covariates.

We used contact testing data and logistic regression to estimate the relationship between index case symptom status and PCR Ct values/viral loads, and positive results in PCR/LFD-tested contacts. We followed a similar approach to previous analyses[13,14] adjusting for multiple index case and contact factors (see Supplement). We used index case-contact pairs plausibly related by transmission to estimate the proportion of infectious index cases potentially detected by LFDs (see Supplement).

### Ethics

Public Health England’s Research Ethics (PHEREG) provided approval for the studies as Service Evaluation and Ongoing Evaluation. This was reviewed and approved under REGG R and D 438 (see Supplement for further details).

### Role of the funding source

LFD evaluations were commissioned by the UK Health Security Agency (UKHSA) and the Department of Health and Social Care (DHSC). Data were analysed by an independent team at the University of Oxford. All authors, including those from the UKHSA and DHSC, contributed to writing the report.

## Results

83,280 paired LFD and PCR tests were performed. 7898 were excluded: 23 were performed with an LFD manufacturer other than the three evaluated, 6389 were tested with endpoint PCR which does not produce comparable estimates of viral load [16] or an unknown PCR type rather than real-time PCR, 46 where the PCR testing was performed at laboratories not participating in the study, 1272 with a void PCR result, 21 participants withdrew from the study, and 147 had void LFD results. This left 75,382 pairs of samples available for analysis performed between 04-November-2020 and 21-March-2022, including 4131 (5.5%) PCR-positive samples (Figure 1). Tests were performed across multiple settings: pre-deployment evaluation (PCR-positive/n 1123/6759, 16.6%), community-based testing (2381/32,266, 7.4%), healthcare workers (28/1587, 1.8%), schools (89/12,397, 0.7%), workplaces (7/5700, 0.1%), universities (38/6456, 0.6%), and in specifically targeted unrepresented groups (465/10,217, 4.6%) (Figure S1).

**Figure 1.**
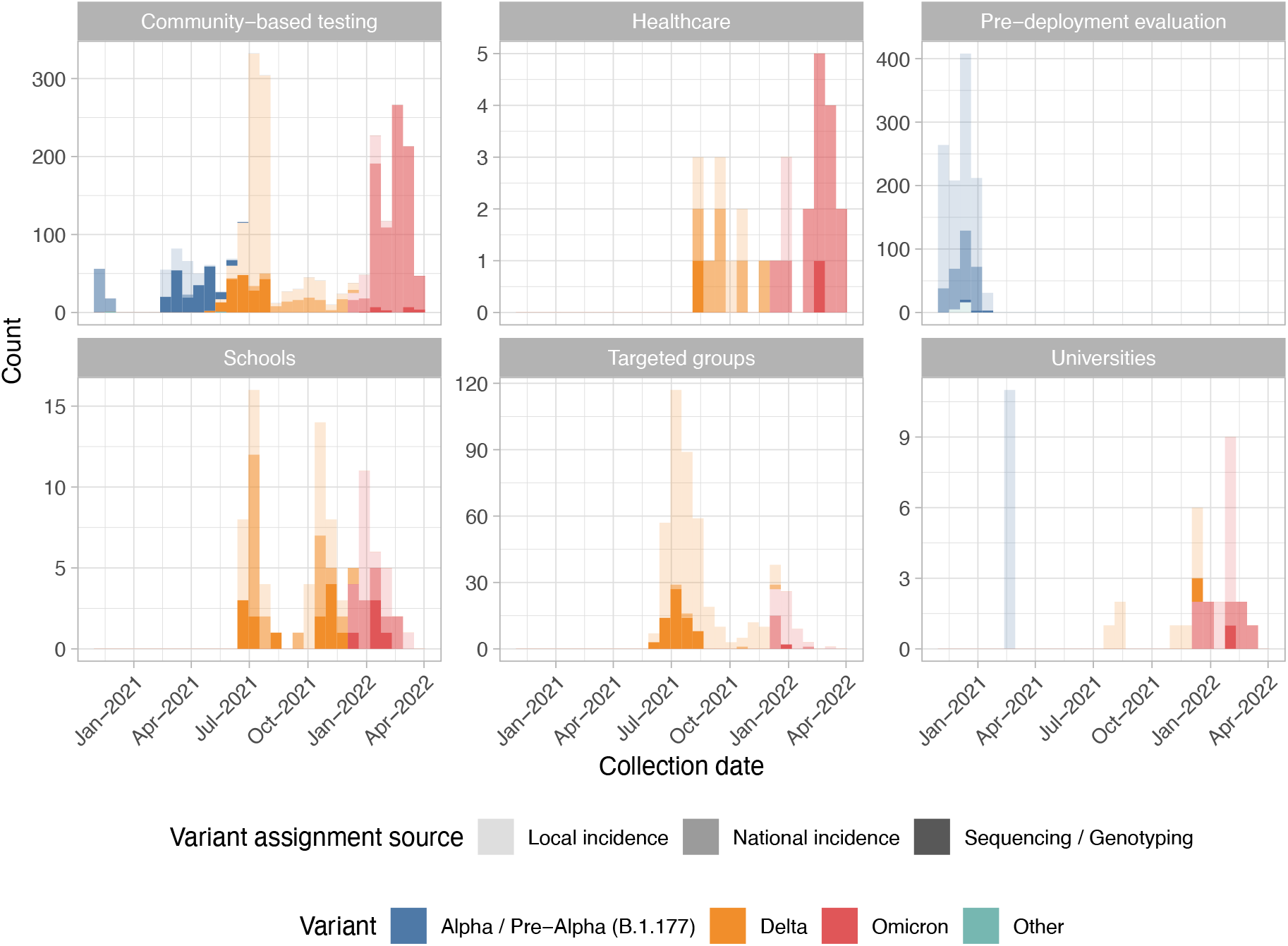
SARS-CoV-2 PCR-positive lateral flow device evaluation samples by study setting and variant. The source of the assigned variant is also shown. Seven PCR-positive results from workplaces are not shown.

### Lateral flow device sensitivity

The overall sensitivity of LFDs relative to PCR-testing was 63.2% (2609/4131, 95%CI 61.7-64.6%). Sensitivity was 71.6% (95%CI 69.8-73.4%) in unselected community-based testing, which was higher than in pre-deployment testing, 52.8% (49.8-55.8%) (Table 1). Unadjusted sensitivity was 55.7% (53.1-58.2%), 64.0% (61.6-66.4%), and 73.0% (70.2-75.6%) during the Alpha/Pre-alpha, Delta and Omicron periods respectively. Unadjusted sensitivity was higher following vaccination, i.e., 67.6% (63.3-71.7%), and 69.7% (67.3-72.0%), after 1, and 2 or more doses respectively, compared to 57.3% (55.1-59.4%) in unvaccinated participants. Sensitivity was higher in symptomatic participants, 68.7% (66.9-70.4%) than in asymptomatic participants, 52.8% (50.1-55.4%).

**Table 1.** SARS-CoV-2 antigen lateral flow device performance in PCR-positive samples. aOR, adjusted odds ratio; CI, confidence interval.

| Characteristic | Summary |  |  | Univariable |  |  | Multivariable |  |  |
| --- | --- | --- | --- | --- | --- | --- | --- | --- | --- |
|  | LFD negative (%), N = 1,524 | LFD positive (Sensitivity %), N = 2,609 | Sensitivity 95% CI | OR | 95% CI | p-value | aOR | 95% CI | p-value |
| Log<sub>10</sub> viral load | 3.84 (2.48, 5.02) | 6.12 (5.29, 6.80) | 5.9, 6.0 | 2.80 | 2.63, 2.98 | <0.001 | 2.85 | 2.66, 3.06 | <0.001 |
| Lateral flow device |  |  |  |  |  |  |  |  |  |
| Innova | 1,270 (38.7%) | 2,008 (61.3%) | 59.6%, 62.9% |  |  |  |  |  |  |
| Acon | 70 (27.7%) | 183 (72.3%) | 66.4%, 77.8% | 1.65 | 1.24, 2.20 | <0.001 | 1.65 | 0.96, 2.83 | 0.070 |
| Orient Gene | 182 (30.3%) | 418 (69.7%) | 65.8%, 73.3% | 1.45 | 1.20, 1.75 | <0.001 | 1.11 | 0.78, 1.57 | 0.6 |
| Study setting |  |  |  |  |  |  |  |  |  |
| Community-based testing | 676 (28.4%) | 1,705 (71.6%) | 69.8%, 73.4% |  |  |  |  |  |  |
| Healthcare | 10 (35.7%) | 18 (64.3%) | 44.1%, 81.4% | 0.71 | 0.33, 1.55 | 0.4 | 0.60 | 0.14, 2.50 | 0.5 |
| Pre-deployment evaluation | 530 (47.2%) | 593 (52.8%) | 49.8%, 55.8% | 0.44 | 0.38, 0.51 | <0.001 | 0.51 | 0.36, 0.73 | <0.001 |
| Schools | 52 (58.4%) | 37 (41.6%) | 31.2%, 52.5% | 0.28 | 0.18, 0.43 | <0.001 | 0.70 | 0.30, 1.63 | 0.4 |
| Targeted groups | 233 (50.1%) | 232 (49.9%) | 45.3%, 54.5% | 0.39 | 0.32, 0.48 | <0.001 | 0.51 | 0.33, 0.78 | 0.002 |
| Universities | 16 (42.1%) | 22 (57.9%) | 40.8%, 73.7% | 0.55 | 0.28, 1.04 | 0.067 | 0.84 | 0.31, 2.33 | 0.7 |
| Workplaces | 5 (71.4%) | 2 (28.6%) | 3.7%, 71.0% | 0.16 | 0.03, 0.82 | 0.028 | 0.12 | 0.01, 1.86 | 0.13 |
| Age | 34 (25, 47) | 36 (25, 47) | 36, 37 | 1.00 | 1.00, 1.01 | 0.3 | 1.00 | 0.99, 1.00 | 0.4 |
| Unknown | 105 | 151 |  |  |  |  |  |  |  |
| Sex |  |  |  |  |  |  |  |  |  |
| Female | 740 (35.9%) | 1,323 (64.1%) | 62.0%, 66.2% |  |  |  |  |  |  |
| Male | 680 (37.4%) | 1,139 (62.6%) | 60.3%, 64.8% | 0.94 | 0.82, 1.07 | 0.3 | 0.89 | 0.75, 1.07 | 0.2 |
| Unknown | 102 | 147 |  |  |  |  |  |  |  |
| Assistance |  |  |  |  |  |  |  |  |  |
| Assisted | 602 (47.5%) | 665 (52.5%) | 49.7%, 55.3% |  |  |  |  |  |  |
| Self | 920 (32.1%) | 1,944 (67.9%) | 66.1%, 69.6% | 1.91 | 1.67, 2.19 | <0.001 | 0.79 | 0.60, 1.05 | 0.11 |
| Symptom status |  |  |  |  |  |  |  |  |  |
| Asymptomatic | 648 (47.2%) | 724 (52.8%) | 50.1%, 55.4% |  |  |  |  |  |  |
| Symptomatic | 847 (31.3%) | 1,859 (68.7%) | 66.9%, 70.4% | 1.96 | 1.72, 2.25 | <0.001 | 1.63 | 1.30, 2.04 | <0.001 |
| Unknown | 27 | 26 |  |  |  |  |  |  |  |
| Vaccination status |  |  |  |  |  |  |  |  |  |
| Unvaccinated | 874 (42.7%) | 1,171 (57.3%) | 55.1%, 59.4% |  |  |  |  |  |  |
| One dose | 163 (32.4%) | 340 (67.6%) | 63.3%, 71.7% | 1.56 | 1.27, 1.91 | <0.001 | 1.13 | 0.81, 1.57 | 0.5 |
| Two or more doses | 471 (30.3%) | 1,083 (69.7%) | 67.3%, 72.0% | 1.72 | 1.49, 1.97 | <0.001 | 1.13 | 0.81, 1.59 | 0.5 |
| Unknown | 14 | 15 |  |  |  |  |  |  |  |
| Variant |  |  |  |  |  |  |  |  |  |
| Alpha / Pre-Alpha (B.1.177) | 674 (44.3%) | 846 (55.7%) | 53.1%, 58.2% |  |  |  |  |  |  |
| Delta | 545 (36.0%) | 970 (64.0%) | 61.6%, 66.4% | 1.42 | 1.23, 1.64 | <0.001 | 1.00 | 0.69, 1.45 | >0.9 |
| Omicron | 290 (27.0%) | 783 (73.0%) | 70.2%, 75.6% | 2.15 | 1.82, 2.55 | <0.001 | 1.63 | 1.02, 2.59 | 0.042 |
| Other | 13 (56.5%) | 10 (43.5%) | 23.2%, 65.5% | 0.61 | 0.27, 1.41 | 0.2 | 0.43 | 0.15, 1.23 | 0.12 |

After adjustment for all other factors, increased viral load was independently associated with a positive LFD result (adjusted odds ratio, aOR=2.85 [95%CI 2.66-3.06] per log10 copies/mL higher), but there was no evidence of a difference in sensitivity between the Innova, Acon, and Orient Gene LFDs (Figure 2, Figure S2). Using the Innova LFD as an example, predicted sensitivity (95%CI) for detecting Omicron infection in community-based symptomatic testing was 28.9% (20.8-38.7%), 77.6% (69.6-83.9%), and 96.7% (95.0-97.9%) at viral loads of 10^3^, 10^5^, and 10^7^ copies/mL respectively. Corresponding values for asymptomatic testing were 19.9% (13.6-28.2%), 67.9% (57.9-76.5%), and 94.7% (91.9-96.6%) respectively.

**Figure 2.**
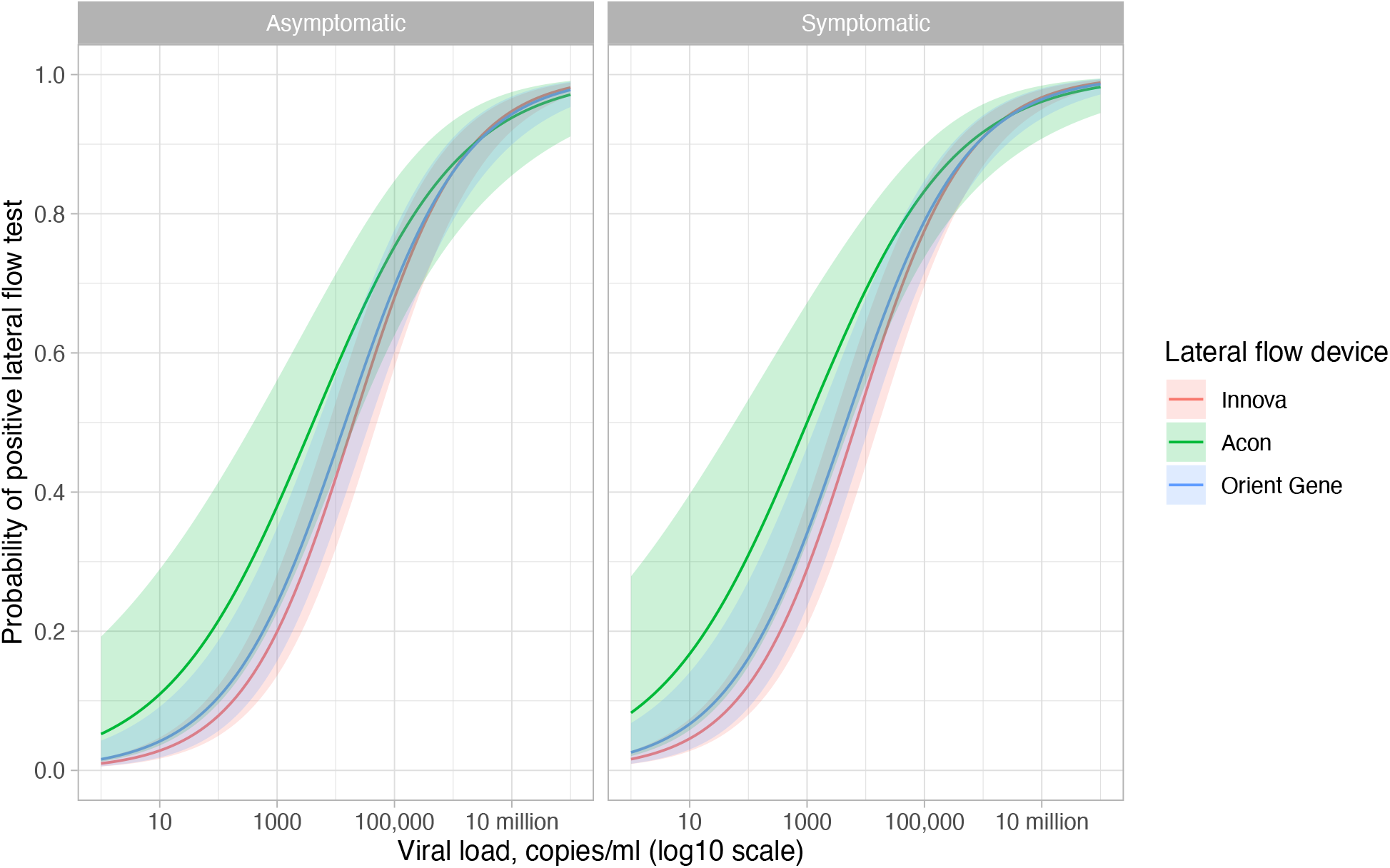
Sensitivity by viral load of SARS-CoV-2 by lateral flow device and patient symptoms. The model fitted adjusts for test setting (predictions are shown for community-based testing), assistance performing the test (self-performed), vaccination status (unvaccinated), and variant (Alpha/Pre-Alpha(B1.1.177)). In addition to the model shown in Table 1 an interaction term between viral load and lateral flow device is included to allow the shape of the curves plotted to vary by device. See Figure S2 for a comparison with observed data.

Compared to community-based testing, sensitivity was independently lower in pre-deployment testing (aOR=0.51 [0.36-0.73]), and in targeted groups (aOR=0.51 [0.33-0.78]). Symptomatic participants were independently more likely to test positive (aOR=1.63 [1.30-2.04]), but there was no evidence of a difference in LFD sensitivity with and without assistance performing the test. After adjusting for other factors there was no evidence that vaccination status, age or sex were associated with LFD results. Similarly, there was no evidence that LFD sensitivity differed with Delta infections compared to Alpha/pre-Alpha infections, but Omicron infections were more likely to be LFD positive than Alpha/pre-Alpha infections (aOR=1.63 [1.02-2.59]).

### Lateral flow device specificity

The overall specificity of LFDs was 99.71% (71,041/71251, 95%CI 99.66-99.74%) and was ≥99.4% across all study settings (Table 2). In a multivariable model, compared to the Innova LFD, Acon devices were associated with 2-times more false-positive results (aOR=2.00 [95%CI 1.21-3.29]), with moderate evidence Orient Gene devices were associated with fewer false-positive results (aOR=0.49 [0.23-1.04]). Compared to community-based testing there were 67% fewer false-positive results in schools (aOR=0.33 [0.16-0.68]) and likely also fewer in universities (aOR=0.34 [0.10-1.16]), whereas false-positive results were 2-times more common in pre-deployment testing (aOR=2.09 [1.15-3.82]). Self-performed tests resulted in >2.5-times more false-positives than assisted tests (aOR=2.61 [1.46-3.13]). False-positive results were 2-times more common in symptomatic compared to asymptomatic participants (aOR=2.15 [1.48-3.13]). There was no evidence of a difference in false-positive results by vaccination status, age or sex. False-positive tests were more common during the Delta (aOR=2.68 [1.43-5.00]) and Omicron (aOR=5.47 [2.54-11.8]) periods compared to the Alpha/Pre-Alpha (B.1.1.177) period.

**Table 2.** SARS-CoV-2 antigen lateral flow device performance in PCR-negative samples. The logistic regression analysis shows odds ratios for a positive test, i.e. higher odds ratios indicate lower specificity. aOR, adjusted odds ratio; CI, confidence interval.

| Characteristic | LFD negative (Specificity %), N = 76,780 | Summary |  | OR | Univariable |  | aOR | Multivariable |  |
| --- | --- | --- | --- | --- | --- | --- | --- | --- | --- |
|  |  | Specificity 95% CI | LFD positive (%), N = 231 |  | 95% CI | p-value |  | 95% CI | p-value |
| LFD device |  |  |  |  |  |  |  |  |  |
| Innova | 59,884 (99.7%) | 99.7%, 99.8% | 156 (0.3%) |  |  |  |  |  |  |
| Acon | 5,712 (99.3%) | 99.0%, 99.5% | 43 (0.7%) | 2.62 | 1.86, 3.67 | <0.001 | 2.00 | 1.21, 3.29 | 0.006 |
| Orient Gene | 5,445 (99.8%) | 99.6%, 99.9% | 11 (0.2%) | 0.70 | 0.38, 1.29 | 0.3 | 0.49 | 0.23, 1.04 | 0.065 |
| Study setting |  |  |  |  |  |  |  |  |  |
| Community-based testing | 29,755 (99.6%) | 99.5%, 99.6% | 130 (0.4%) |  |  |  |  |  |  |
| Healthcare | 1,552 (99.6%) | 99.1%, 99.8% | 7 (0.4%) | 1.03 | 0.48, 2.21 | >0.9 | 1.33 | 0.48, 3.71 | 0.6 |
| Pre-deployment evaluation | 5,602 (99.4%) | 99.2%, 99.6% | 34 (0.6%) | 1.39 | 0.95, 2.03 | 0.089 | 2.09 | 1.15, 3.82 | 0.016 |
| Schools | 12,292 (99.9%) | 99.8%, 99.9% | 16 (0.1%) | 0.30 | 0.18, 0.50 | <0.001 | 0.33 | 0.16, 0.68 | 0.003 |
| Targeted groups | 9,732 (99.8%) | 99.7%, 99.9% | 20 (0.2%) | 0.47 | 0.29, 0.75 | 0.002 | 0.97 | 0.45, 2.09 | >0.9 |
| Universities | 6,415 (100.0%) | 99.9%, 100.0% | 3 (0.0%) | 0.11 | 0.03, 0.34 | <0.001 | 0.34 | 0.10, 1.16 | 0.085 |
| Workplaces | 5,693 (100.0%) | 99.9%, 100.0% | 0 (0.0%) |  |  |  |  |  |  |
| Assistance |  |  |  |  |  |  |  |  |  |
| Assisted | 30,095 (99.9%) | 99.8%, 99.9% | 41 (0.1%) |  |  |  |  |  |  |
| Self | 40,946 (99.6%) | 99.5%, 99.6% | 169 (0.4%) | 2.46 | 1.75, 3.46 | <0.001 | 2.61 | 1.46, 4.67 | 0.001 |
| Symptom status |  |  |  |  |  |  |  |  |  |
| Asymptomatic | 56,766 (99.8%) | 99.7%, 99.8% | 119 (0.2%) |  |  |  |  |  |  |
| Symptomatic | 10,640 (99.2%) | 99.0%, 99.3% | 87 (0.8%) | 3.65 | 2.77, 4.82 | <0.001 | 2.15 | 1.48, 3.13 | <0.001 |
| Unknown | 3,635 |  | 4 |  |  |  |  |  |  |
| Vaccination status |  |  |  |  |  |  |  |  |  |
| Unvaccinated | 24,677 (99.7%) | 99.6%, 99.8% | 73 (0.3%) |  |  |  |  |  |  |
| One dose | 12,263 (99.8%) | 99.7%, 99.9% | 27 (0.2%) | 0.89 | 0.57, 1.39 | 0.6 | 0.76 | 0.43, 1.35 | 0.4 |
| Two or more doses | 32,681 (99.7%) | 99.6%, 99.7% | 110 (0.3%) | 1.08 | 0.81, 1.46 | 0.6 | 0.81 | 0.47, 1.40 | 0.4 |
| Unknown | 1,420 |  | 0 |  |  |  |  |  |  |
| Dominant circulating variant |  |  |  |  |  |  |  |  |  |
| Alpha / Pre-Alpha (B.1.177) | 24,440 (99.8%) | 99.7%, 99.8% | 53 (0.2%) |  |  |  |  |  |  |
| Delta | 39,297 (99.7%) | 99.7%, 99.8% | 114 (0.3%) | 1.16 | 0.84, 1.61 | 0.4 | 2.68 | 1.43, 5.00 | 0.002 |
| Omicron | 7,098 (99.4%) | 99.2%, 99.6% | 41 (0.6%) | 2.21 | 1.47, 3.33 | <0.001 | 5.47 | 2.54, 11.8 | <0.001 |
| Other | 206 (99.0%) | 96.6%, 99.9% | 2 (1.0%) | 3.78 | 0.91, 15.6 | 0.066 | 2.52 | 0.58, 10.9 | 0.2 |
| Age |  |  |  | 0.99 | 0.98, 1.00 | 0.11 | 0.99 | 0.98, 1.00 | 0.2 |
| Unknown |  |  |  |  |  |  |  |  |  |
| Sex |  |  |  |  |  |  |  |  |  |
| Female |  |  |  | 0.99 | 0.74, 1.33 | >0.9 | 1.06 | 0.78, 1.43 | 0.7 |
| Male |  |  |  |  |  |  |  |  |  |

### Proportion of infectious individuals detected by lateral flow devices

Between 01-January-2021 and 11-January-2022 6,263,786 contacts of PCR-positive SARS-CoV-2 index cases were identified, 1,173,643 (18.7%) underwent a PCR or reported LFD in the 1 to 10 days following the index case’s PCR test, 377,151 (6.0%) tested positive. Contacts were linked to 347,374 unique index cases, with 322,416 (92.8%) index cases linked to a single positive contact, 21,321 (6.1%) to two positive contacts and 3637 (1.0%) to ≥3 (Figure S3).

As described previously,[13] tested contacts of index cases with higher viral loads (lower Ct values) were more likely to test positive. Contacts of symptomatic index cases were more likely to test positive than those of asymptomatic index cases, even at a given viral load (Figure 3). Additionally, viral loads in asymptomatic index cases were lower than those in symptomatic index cases throughout the Alpha, Delta and Omicron periods (Figure S4).

**Figure 3.**
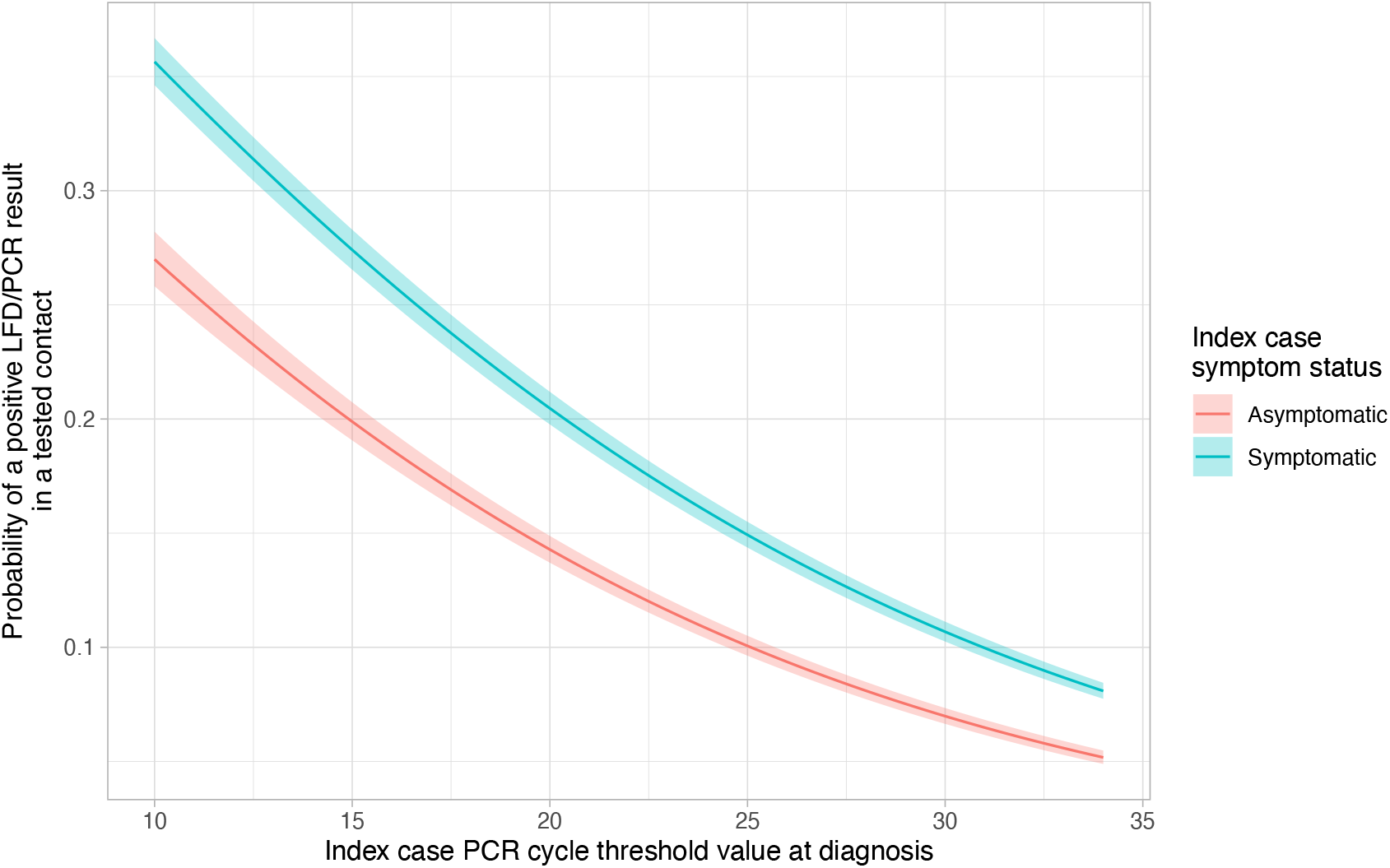
Relationships between index case PCR cycle threshold (Ct) value and probability of a tested contact being PCR/LFD-positive, by index case symptom status. Contacts of asymptomatic index cases are 0.76 times as likely to test positive as contacts of symptomatic index cases at a Ct value of 10, 0.70 times at a Ct value of 20 and 0.65 times at a Ct value of 30. Model adjusted for index case age (set to 40 years), index case sex (set to female), index case vaccination status (set to boosted), contact event type (set to household or accommodation), contact age (set to 40 years), contact sex (set to female) and test date (set to 01 July 2021). There was no evidence that fitting an interaction between index case symptom status and Ct values improved model fit.

79.4% (95%CI 68.6-81.3%) of index cases with ≥1 PCR/LFD-positive contact were estimated to have been detectable by an LFD. Similar detection rates were seen in index cases with 1, 2 or ≥3 PCR/LFD-positive contacts, 79.4% (68.5-81.2%), 80.2% (69.1-81.7%), 79.6% (68.1-81.2%), respectively. Amongst index cases with ≥1 PCR/LFD-positive contact, 20,712 were recorded as asymptomatic and 326,662 as symptomatic; the proportion of index cases estimated as detectable by LFD was higher with symptoms (80.8% [95%CI 69.3-82.1%]) than without (59.1% [55.2-69.0%]).

The estimated proportion of index cases detected by LFD was similar over time and across the Alpha, Delta, and Omicron periods for symptomatic cases (Figure 4, Table 3). For asymptomatic index cases, the estimated percentage detected by LFDs was consistently lower than for symptomatic index cases, but within asymptomatic index cases higher during the Omicron period compared to the Alpha and Delta periods. This reflects the increase in measured sensitivity of LFDs during the Omicron period rather than a change in the distribution of viral loads (Figure S4).

**Figure 4.**
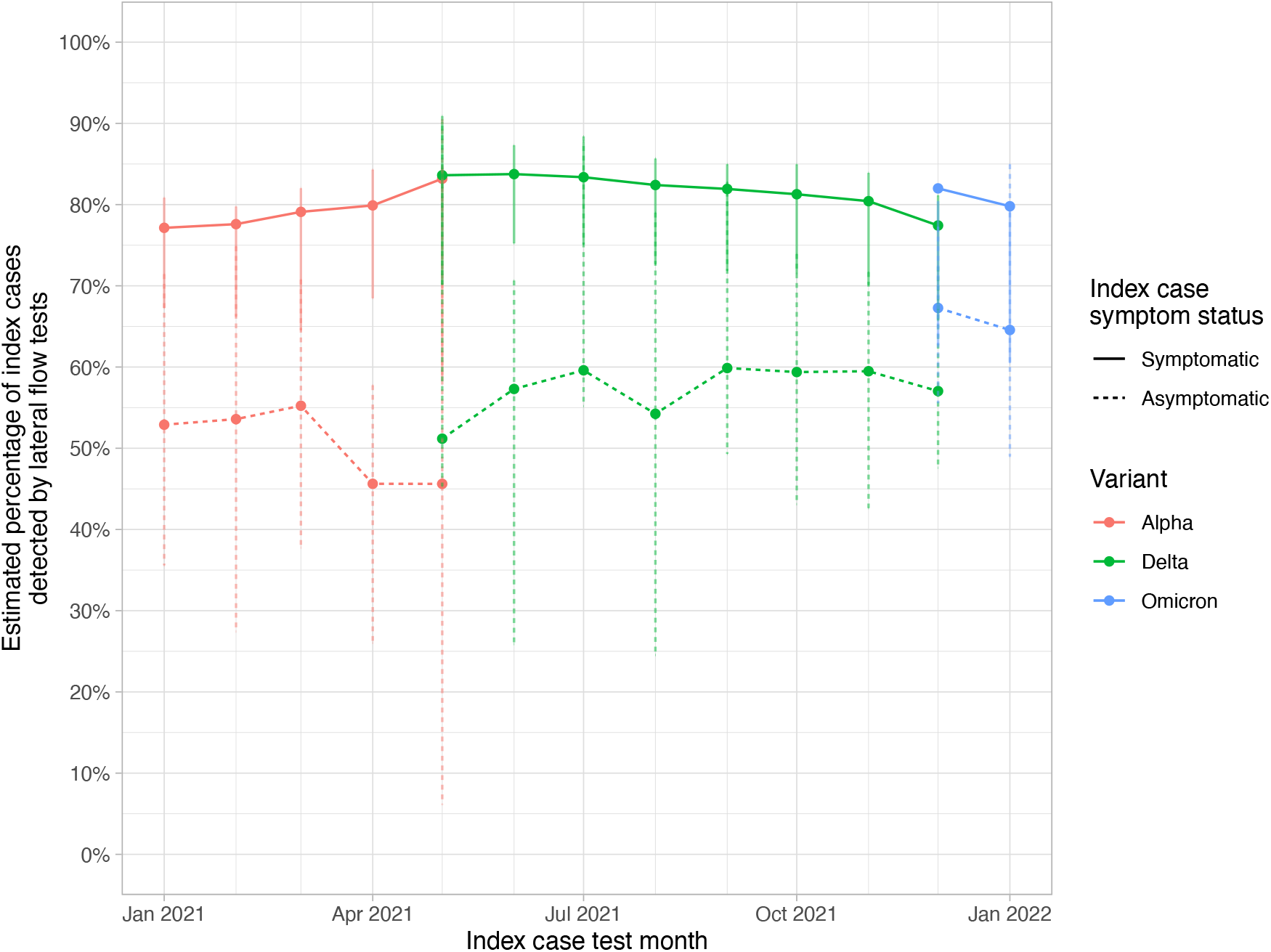
Percentage of index cases estimated to be detected by lateral flow device amongst case-contact pairs with probable SARS-CoV-2 transmission. Data are plotted aggregated by month with lines coloured by estimated variant, and the line type indicated index case symptom status. Error bars indicate 95% confidence intervals calculated by non-parametric bootstrap.

**Table 3.** Percentage of index cases estimated to be detected by lateral flow device amongst case-contact pairs with probable SARS-CoV-2 transmission.

| **Estimated variant** | **Index case symptom status** | **Index cases** | **Percentage of index cases estimated to be detected by lateral flow devices** | **95% confidence interval** |
| --- | --- | --- | --- | --- |
| Alpha | Symptomatic | 60,322 | 77.6% | 64.5-83.3% |
| Delta | Symptomatic | 198,707 | 81.5% | 69.6-86.9% |
| Omicron | Symptomatic | 67,633 | 81.3% | 61.3-79.9% |
| Alpha | Asymptomatic | 3,590 | 52.5% | 17.7-72.3% |
| Delta | Asymptomatic | 13,153 | 58.6% | 37.3-86.5% |
| Omicron | Asymptomatic | 3,969 | 66.4% | 51.7-81.0% |

## Discussion

In a national LFD evaluation program, the sensitivity of LFDs compared with PCR was 71.6% (95%CI 69.8-73.4%) in 2381 paired PCR-positive samples obtained in routine community-based testing. There was no evidence that LFD sensitivity or specificity was independently associated with vaccination status, age or sex. Performance was similar during the Alpha/Pre-Alpha and Delta periods, with an increase in sensitivity during the Omicron period. This potentially reflects either intrinsic changes in the SARS-CoV-2 virus or changes in testing proficiency and behaviour around testing over time but is in contrast to other reports of lower sensitivity with Omicron,[17,18] while others have reported similar performance with Omicron vs. Delta.[19] Conversely, although specificity remained >99%, it declined over time with successive variants.

Symptomatic individuals had higher viral loads in the LFD evaluations. However, even having adjusted for this, the presence of fever, cough or anosmia/ageusia were associated with increased LFD sensitivity at a given viral load compared to without these cardinal symptoms. This may reflect different ratios of antigen to RNA with different symptom statuses and day of infection. With a one-off screen asymptomatic individuals may be detected later in their infections where residual RNA may be present with less antigen compared to earlier in infection, when most symptomatic cases were tested. Serial testing by PCR and quantitative antigen tests could help investigate this further.

There was no evidence of a difference in overall sensitivity between the different LFD manufacturers tested, potentially reflecting limited power as most paired tests were performed with the Innova LFD. However, when considering sensitivity by viral load, there was some evidence that the Acon device was more sensitive at lower viral loads, but this was also the device with the lowest specificity, likely reflecting a trade off in how devices are calibrated.

LFD sensitivity increased with viral load, such that those with higher viral loads, i.e. those who were more infectious, were more likely to detected by LFDs.[20–23] Amongst case-contact pairs where transmission was plausible, the percentage of index cases detected by LFDs was 79.4%, similar to previous estimates with the same LFD device.[14] This is higher than the overall sensitivity of LFDs at 71.6% in community testing. However, we show that the potential for LFDs to detect infectious asymptomatic index cases was lower than for symptomatic index cases, at 59.1% vs. 80.8%. This is the result of a combination of reduced sensitivity at a given viral load and reduced viral loads in asymptomatic infections in the contact testing data. However, asymptomatic individuals are less infectious than symptomatic individuals at a given viral load, and the lower viral loads in asymptomatic individuals are also associated with less transmission.[13] It is also worth noting that asymptomatic index cases are likely to be under ascertained in national contact tracing data, especially during periods when asymptomatic screening was uncommon.

Taken together our findings provide support for the use of LFDs as a mechanism to detect potentially infectious individuals and reduce transmission, particularly as these tests perform best in those most likely to be infectious, i.e., those who are symptomatic and/or with high viral loads. However, we show, alongside other studies, that the performance of these tests in asymptomatic individuals may be lower than is generally understood.[4,22,24] LFDs have been deployed in several settings including population-wide mass testing of asymptomatic individuals. In this setting a combination of population prevalence, lower viral loads in asymptomatic individuals, and lower onward transmission at a given viral load compared to symptomatic cases mean that even with perfect LFD sensitivity likely several thousand asymptomatic people need to be tested to detect one case who would otherwise go on to transmit (Table S2). As the sensitivity of LFDs falls, more individuals need to be tested to detect one potentially transmitting case. If false-negative results do not change behaviour, then reduced LFD sensitivity acts simply to make testing programmes less efficient and more costly per transmission averted, but the use of LFD is still potentially effective at reducing the spread of infections. Hence, where people plan to participate in activities anyway then LFDs may act to reduce risk, even if imperfectly. Another setting where risk reduction may be important is where the consequences of transmission are high and those tested are likely to maintain transmission precautions with a negative result, e.g. in healthcare or social care. However, some caution is required if LFDs are used by individuals to relax adhering to transmission precautions.[25,26] Here it is possible that false-negative results with a poorly sensitive test could lead to additional transmission. This possibility needs to be considered when assessing the implementation and messaging around potential asymptomatic screening programs.

This study has several limitations regarding the evaluation of LFD performance. As testing was undertaken in real-world settings, the primary diagnostic test was performed first, rather than randomising the order of swabs taken for LFDs and PCR. A range of different PCR tests were used as a reference standard, however calibrants were used to convert Ct values across assays to estimated viral loads in common units. The calibration approach adopted using a known reference standard was deployable across all test sites, but more accurate approaches for quantifying viral load including droplet digital PCR exist. It should also be noted that viral loads estimated in the samples only approximate viral loads in the participants given respiratory samples are based on swabs rather than direct sampling of a body fluid. Although we show that asymptomatic individuals were less likely to be detected by LFDs, in part this may reflect the performance of a single test which may have captured some infections several days after they started. It is possible that regular asymptomatic testing would have improved performance, as this may catch incident infections earlier in their course. We also did not make use of digital LFD readers which may increase accuracy and have since been widely deployed in the UK.[27,28]

The transmission analysis also has limitations. The case-contact pairs identified rely on both index case and contacts participating in SARS-CoV-2 testing, with several demographic, socioeconomic, and behavioural factors potentially affecting test-seeking. One particular concern is that during the period of the study PCR testing was provided for symptomatic individuals or following a positive LFD test. However, some asymptomatic individuals also sought PCR testing for other reasons, e.g. following contact events. It is therefore possible that case-contact pairs involving an asymptomatic case are enriched for pairs with contact with a third party who is the true source of both infections. In this case the properties of the asymptomatic cases are not what determined transmission.

In summary, LFDs have remained able to detect most SARS-CoV-2 infections throughout the roll-out of vaccination and with several different viral variants. Although on-going monitoring of performance with new variants is required while tests are used, it is reassuring that LFDs are probably likely to remain able to detect future variants. LFDs potentially detect most infections that have the potential to transmit to others, however performance is lower in asymptomatic compared to symptomatic individuals and this needs to be considered when designing testing programs.

## Data Availability

Applications to use the data in this study can be made to NHS Digital's Data Access Request Service, please see https://digital.nhs.uk/services/data-access-request-service-dars for more details.

## Transparency declaration

DWE has received lecture fees from Gilead outside the submitted work. No other author has a conflict of interest to declare.

## Funding

Supported by the UK Government Department of Health and Social Care; the National Institute for Health Research (NIHR) Health Protection Research Unit in Healthcare Associated Infections and Antimicrobial Resistance, University of Oxford, in partnership with Public Health England (NIHR200915); and the University of Oxford NIHR Biomedical Research Centre. DWE is a Robertson Foundation Fellow.

## Notes

### Author Declarations

Public Health England's Research Ethics (PHEREG) provided approval for the studies as Service Evaluation and Ongoing Evaluation. This was reviewed and approved under REGG R and D 438.

## References

1. Mahase E. Covid-19: UK regulator approves lateral flow test for home use despite accuracy concerns. Bmj. 2020;371: m4950. doi:10.1136/bmj.m4950

2. NHS England. Coronavirus “NHS England and NHS Improvement rollout of lateral flow devices for asymptomatic staff testing for SARS CoV-2 (phase 2: trusts). 16 Nov 2020 [cited 13 Nov 2022]. Available: https://www.england.nhs.uk/coronavirus/documents/nhs-england-and-nhs-improvement-rollout-of-lateral-flow-devices-for-asymptomatic-staff-testing-for-sars-cov-2-phase-2-trusts/

3. Deeks J, Raffle A, Gill M. Covid-19: government must urgently rethink lateral flow test roll out - The BMJ. 12 Jan 2021 [cited 13 Nov 2022]. Available: https://blogs.bmj.com/bmj/2021/01/12/covid-19-government-must-urgently-rethink-lateral-flow-test-roll-out/

4. Dinnes J, Deeks JJ, Berhane S, Taylor M, Adriano A, Davenport C, et al. Rapid, point-of-care antigen tests for diagnosis of SARS-CoV-2 infection. Cochrane Db Syst Rev. 2021;2022: CD013705. doi:10.1002/14651858.cd013705.pub2

5. Mistry DA, Wang JY, Moeser M-E, Starkey T, Lee LYW. A systematic review of the sensitivity and specificity of lateral flow devices in the detection of SARS-CoV-2. Bmc Infect Dis. 2021;21: 828. doi:10.1186/s12879-021-06528-3

6. Department of Health and Social Care. Asymptomatic testing for SARS-CoV-2 using antigen-detecting lateral flow devices. 2021. Available: https://assets.publishing.service.gov.uk/government/uploads/system/uploads/attachment_data/file/999866/asymptomatic-testing-for-SARS-CoV-2-using-antigen-detecting-lateral-flow-devices-evidence-from-performance-data-Oct-2020-to-May-2021.pdf

7. Deeks JJ, Raffle AE. Lateral flow tests cannot rule out SARS-CoV-2 infection. Bmj. 2020;371: m4787. doi:10.1136/bmj.m4787

8. Howard J, Huang A, Li Z, Tufekci Z, Zdimal V, Westhuizen H-M van der, et al. An evidence review of face masks against COVID-19. Proc National Acad Sci. 2021;118: e2014564118. doi:10.1073/pnas.2014564118

9. Morris DH, Rossine FW, Plotkin JB, Levin SA. Optimal, near-optimal, and robust epidemic control. Commun Phys. 2021;4: 78. doi:10.1038/s42005-021-00570-y

10. Fisman DN, Greer AL, Tuite AR. Bidirectional impact of imperfect mask use on reproduction number of COVID-19: A next generation matrix approach. Infect Dis Model. 2020;5: 405–408. doi:10.1016/j.idm.2020.06.004

11. Quilty BJ, Clifford S, Hellewell J, Russell TW, Kucharski AJ, Flasche S, et al. Quarantine and testing strategies in contact tracing for SARS-CoV-2: a modelling study. Lancet Public Heal. 2021;6: e175–e183. doi:10.1016/s2468-2667(20)30308-x

12. Petersen I, Crozier A, Buchan I, Mina MJ, Bartlett JW. Recalibrating SARS-CoV-2 Antigen Rapid Lateral Flow Test Relative Sensitivity from Validation Studies to Absolute Sensitivity for Indicating Individuals Shedding Transmissible Virus. Clin Epidemiology. 2021;13: 935–940. doi:10.2147/clep.s311977

13. Eyre DW, Taylor D, Purver M, Chapman D, Fowler T, Pouwels KB, et al. Effect of Covid-19 Vaccination on Transmission of Alpha and Delta Variants. New Engl J Med. 2022;386: 744–756. doi:10.1056/nejmoa2116597

14. Lee LYW, Rozmanowski S, Pang M, Charlett A, Anderson C, Hughes GJ, et al. SARS-CoV-2 infectivity by viral load, S gene variants and demographic factors and the utility of lateral flow devices to prevent transmission. Clin Infect Dis. 2021; ciab421-. doi:10.1093/cid/ciab421

15. University of Liverpool. Covid-SMART Asymptomatic Testing Pilot in Liverpool City Region: Quantitative Evaluation. In: 20/12/21 [Internet]. [cited 11 Nov 2022]. Available: https://www.liverpool.ac.uk/media/livacuk/coronavirus/Liverpool_City_Region_Covid_SMART_Evaluation-Feb.pdf

16. Department of Health and Social Care. Evaluation of endpoint PCR (EPCR) as a central laboratory based diagnostic test technology for SARS-CoV-2. 28 Jan 2021 [cited 11 Nov 2022]. Available: https://www.gov.uk/government/publications/evaluation-of-endpoint-pcr-epcr-as-a-diagnostic-test-technology-for-sars-cov-2/evaluation-of-endpoint-pcr-epcr-as-a-central-laboratory-based-diagnostic-test-technology-for-sars-cov-2

17. Schuit E, Venekamp RP, Hooft L, Veldhuijzen IK, Bijllaardt W van den, Pas SD, et al. Diagnostic accuracy of covid-19 rapid antigen tests with unsupervised self-sampling in people with symptoms in the omicron period: cross sectional study. Bmj. 2022;378: e071215. doi:10.1136/bmj-2022-071215

18. Osterman A, Badell I, Basara E, Stern M, Kriesel F, Eletreby M, et al. Impaired detection of omicron by SARS-CoV-2 rapid antigen tests. Med Microbiol Immun. 2022;211: 105–117. doi:10.1007/s00430-022-00730-z

19. Soni A, Herbert C, Filippaios A, Broach J, Colubri A, Fahey N, et al. Comparison of Rapid Antigen Tests’ Performance Between Delta and Omicron Variants of SARS-CoV-2. Ann Intern Med. 2022; M22–0760. doi:10.7326/m22-0760

20. Singanayagam A, Patel M, Charlett A, Bernal JL, Saliba V, Ellis J, et al. Duration of infectiousness and correlation with RT-PCR cycle threshold values in cases of COVID-19, England, January to May 2020. Eurosurveillance. 2020;25: 2001483. doi:10.2807/1560-7917.es.2020.25.32.2001483

21. Landaas ET, Storm ML, Tollånes MC, Barlinn R, Kran A-MB, Bragstad K, et al. Diagnostic performance of a SARS-CoV-2 rapid antigen test in a large, Norwegian cohort. J Clin Virol. 2021;137: 104789. doi:10.1016/j.jcv.2021.104789

22. Parvu V, Gary DS, Mann J, Lin Y-C, Mills D, Cooper L, et al. Factors that Influence the Reported Sensitivity of Rapid Antigen Testing for SARS-CoV-2. Front Microbiol. 2021;12: 714242. doi:10.3389/fmicb.2021.714242

23. Jääskeläinen A, Ahava M, Jokela P, Szirovicza L, Pohjala S, Vapalahti O, et al. Evaluation of three rapid lateral flow antigen detection tests for the diagnosis of SARS-CoV-2 infection. J Clin Virol. 2021;137: 104785. doi:10.1016/j.jcv.2021.104785

24. Tapari A, Braliou GG, Papaefthimiou M, Mavriki H, Kontou PI, Nikolopoulos GK, et al. Performance of Antigen Detection Tests for SARS-CoV-2: A Systematic Review and Meta-Analysis. Diagnostics. 2022;12: 1388. doi:10.3390/diagnostics12061388

25. Deeks JJ, Singanayagam A, Houston H, Sitch AJ, Hakki S, Dunning J, et al. SARS-CoV-2 antigen lateral flow tests for detecting infectious people: linked data analysis. Bmj. 2022;376: e066871. doi:10.1136/bmj-2021-066871

26. García-Fiñana M, Hughes DM, Cheyne CP, Burnside G, Stockbridge M, Fowler TA, et al. Performance of the Innova SARS-CoV-2 antigen rapid lateral flow test in the Liverpool asymptomatic testing pilot: population based cohort study. Bmj. 2021;374: n1637. doi:10.1136/bmj.n1637

27. UK Health Security Agency. LFD Digital Reader. Evaluation of real world deployment: final report. 1 Jul 2022 [cited 13 Nov 2022]. Available: https://assets.publishing.service.gov.uk/government/uploads/system/uploads/attachment_data/file/1115461/LFD-digital-reader-november-2022.pdf

28. Consortium TLA, Beggs AD, Caiado CCS, Branigan M, Lewis-Borman P, Patel N, et al. Machine learning for determining lateral flow device results for testing of SARS-CoV-2 infection in asymptomatic populations. Cell Reports Medicine. 2022;3: 100784. doi:10.1016/j.xcrm.2022.100784

