## Supplementary material for "Performance of antigen lateral flow devices in the United Kingdom during the Alpha, Delta, and Omicron waves of the SARS-CoV-2 pandemic"

### Supplementary Methods

#### Swab details

PCR testing was performed on combined anterior nose and throat swabs, however nose only swabs were considered acceptable if swabbing both the throat and nose was not possible, e.g. in a distressed child. Innova LFDs were performed using a combined anterior nose and throat swab. Acon and Orient Gene LFDs were performed on anterior nose swabs.

#### Statistical analysis

Data were linked within UKHSA systems and subsequently deidentified prior to being extracted for analysis. For LFD performance evaluations the infecting variant in PCR-positive infections was assigned based on sequencing or PCR-based genotyping where available and if not, based on the dominant variant in sequenced samples from the same week in the participant's local region (Lower Tier Local Authority) if >50% of samples were of a single variant and the address of the participant was known. Where the address was unknown if >50% of sequenced samples nationally were of a single variant then this variant was assigned (before 19 May 2021, Alpha/Pre-Alpha; 19 May – 12 December 2021, Delta; after 12 December 2021, Omicron), otherwise the variant was set to unknown. Based on the sequenced or genotyped samples in the dataset (n=517), we estimated the precision of this approach (i.e. percentage of correctly predicted variants) to be larger than 96%. A higher setting of the threshold for the percentage of cases due to a single variant could increase the precision to over 99% but only at the expense of a substantial loss of coverage to 60%. Similarly, to assess the specificity of LFDs according to time epochs defined by the dominant variant, local and national incidence data were used to assign a variant epoch to each sample.

Real time PCR cycle threshold (Ct) values were used to estimate SARS-CoV-2 viral loads in copies/mL using conversion formulae derived for each laboratory using calibrant samples (Qnostics SCV2AQP01 quantitative SARS-CoV-2 standards panel, calibrated in digital droplet PCR copies per mL; Table S1). Sample pairs only tested by endpoint PCR were excluded.

PCR-positive samples were used for analyses of LFD sensitivity. Univariable and multivariable logistic regression was used to model the relationship between LFD positivity and $\log_{10}$ viral load and other covariates. Covariates included LFD device, study setting, assisted vs. self-testing, self-reported symptom status (symptomatic, i.e. fever, cough or anosmia/ageusia, otherwise asymptomatic), vaccination status by number of doses (0, 1, 2, or 3) and viral variant (Alpha [B.1.1.7] / pre-Alpha [B.1.177], Delta [B.1.617.2], Omicron [BA.1 and BA.2]; or Other / Unknown). PCR-negative samples were used to analyse LFD specificity using univariable and multivariable logistic regression and the same covariates.

We used contact testing data and logistic regression to estimate the relationship between index case symptom status and PCR Ct values/viral loads, and positive results in PCR/LFD-tested contacts. We used the same Ct value to viral load conversions used above (Table S1).

We followed a similar approach to previous analyses[1,2] adjusting for index case age, sex, vaccination status (partial, two doses, boosted), contact event type, contact age, sex, vaccination status, and calendar time (as a proxy for changes with time, incidence, and circulating variants). Natural cubic splines were used to account for non-linearity in continuous variables (4 knots; except calendar time, 8 knots). Pre-specified interactions based on previous analyses [1] were included between contact event type and index case age, contact event type and contact age, index case sex and contact sex, index case age and contact age.

We used index case-contact pairs plausibly related by transmission to estimate the proportion of infectious index cases potentially detected by LFDs. Each index case was included in the analysis only once, however the total number of linked PCR/LFD-positive contacts per index case was also recorded to allow evaluation of whether performance differed in more infectious index cases. We performed a separate analysis by month from 01 January 2021 to 11 January 2022, using data from all index cases with at least one PCR/LFD-positive contact. The analysis was ended on 11 January 2022 as after this date the requirement for positive LFDs to be confirmed by PCR was dropped, meaning both index cases and infected contacts were less well ascertained. To estimate the probability of each index case being LFD-positive, we applied the estimated performance in community testing of the most widely used LFD (Innova), accounting for the symptom status of the index case and the dominant variant at each index case's diagnosis (Alpha, Delta or Omicron). We used non-parametric bootstrap sampling with replacement of index cases (1000 iterations) to estimate 95% confidence intervals. To ensure our estimates reflected the uncertainty in LFD performance estimates we also applied bootstrap sampling to the LFD performance data, re-estimating LFD performance for each iteration.

### Ethics

Within the context of the pandemic public health response and roll out of testing interventions, after review using the Health Research Authority (HRA) tool and further discussions with HRA it was determined that this evaluation would not require HRA research ethics approval. After an initial period, it was determined to gain Public Health England's Research Ethics (PHEREG) approval (then separate to NHS Test and Trace) as service evaluations for subsequent studies to ensure further external scrutiny and assurance on this approach. Approval was obtained for an umbrella framework and associated participant-facing materials for the prospective data collection elements of Service Evaluation and Ongoing Evaluation. This was reviewed and approved under REGG R and D 438.

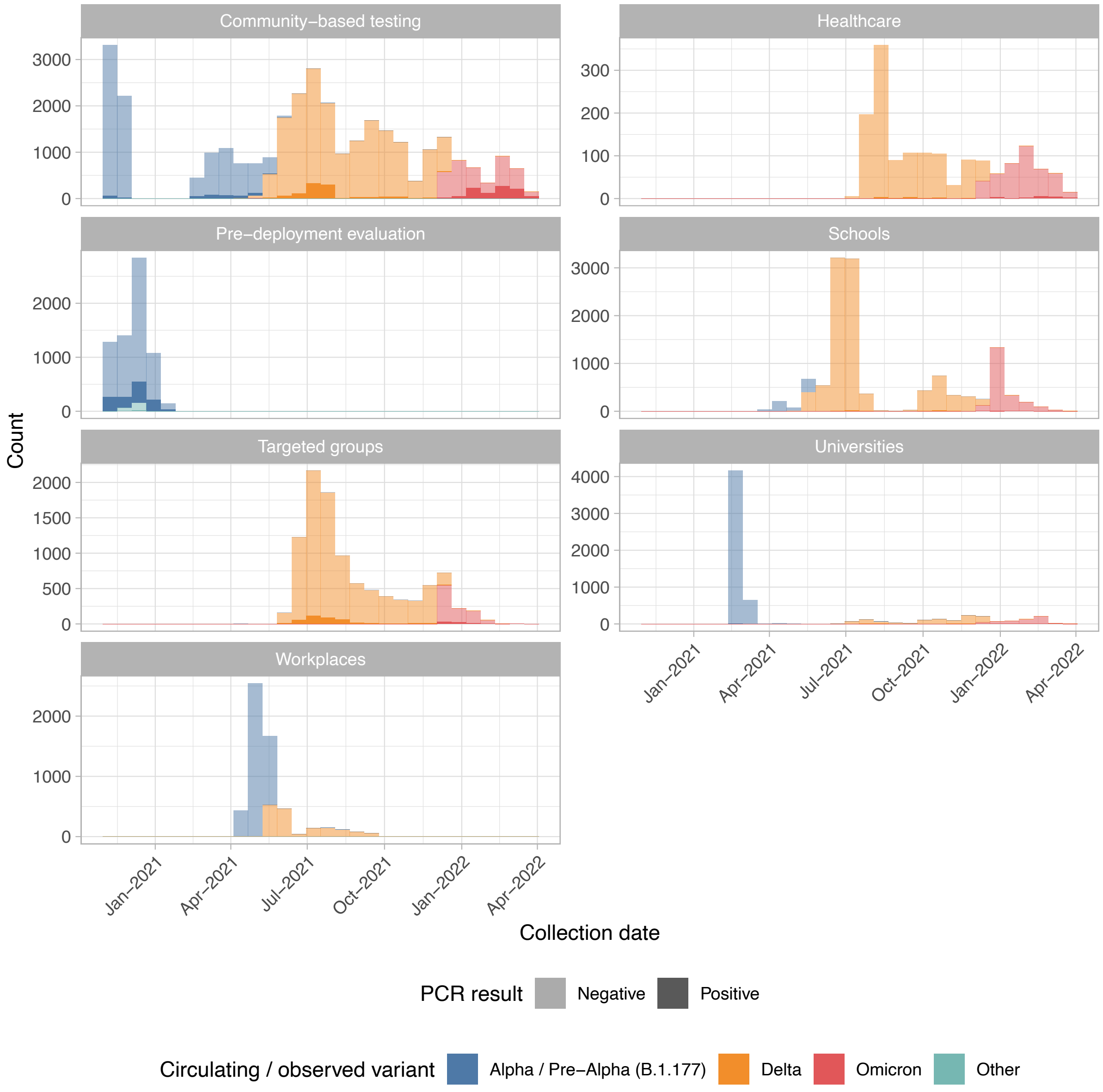


**Figure S1. Lateral flow device evaluation samples by study setting, PCR result and circulating / observed variant**.

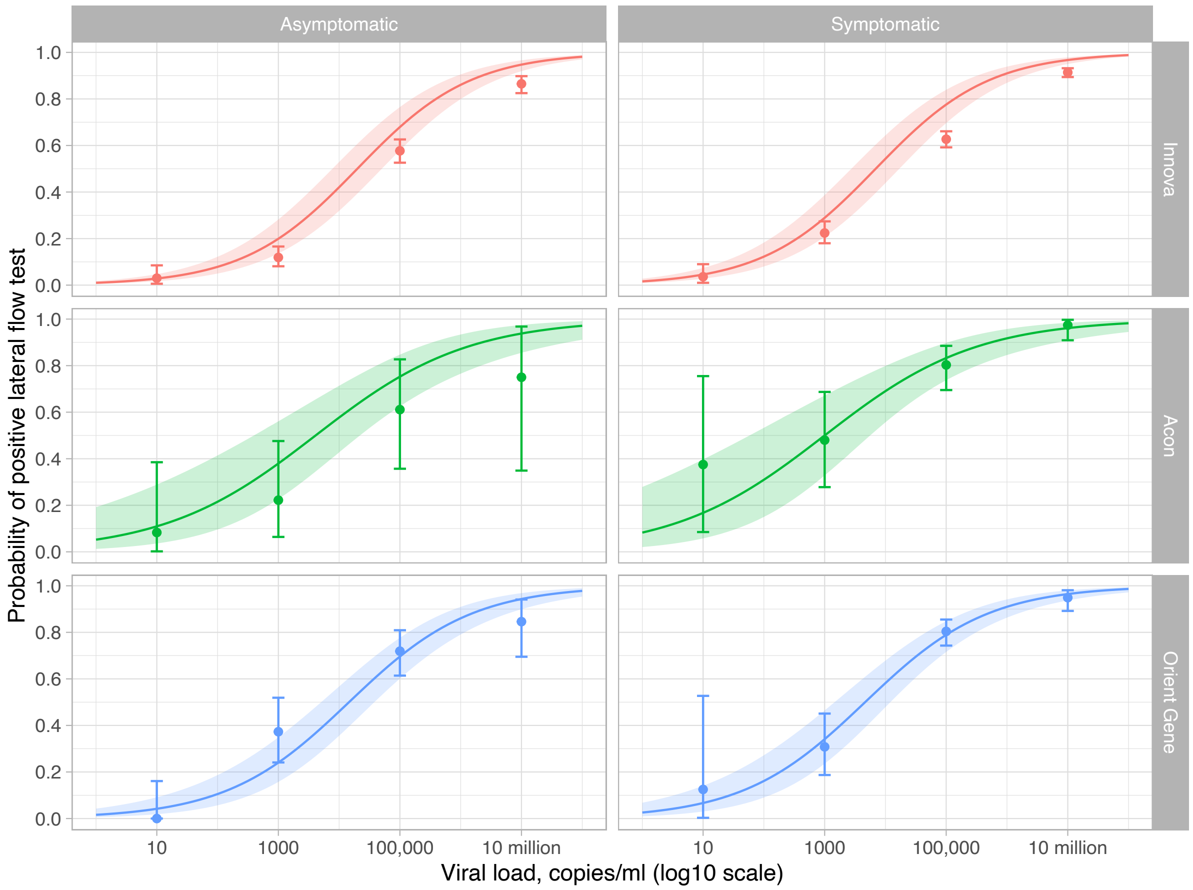


**Figure S2. Sensitivity by viral load of SARS-CoV-2 lateral flow device and patient symptoms**. Points (with error bars indicating exact binomial confidence intervals) are the observed data for <100 copies/ml, 100 to <10,000 copies/ml, 10,000 to <1 million copies/ml and 1 million to <100 million copies/ml. Model estimates shown by the continuous line were obtained from a logistic regression model adjusting viral load, symptom status, lateral flow device, test setting (predictions are shown for community-based testing), assistance performing the test (self-performed), vaccination status (unvaccinated), and variant (Alpha/Pre-Alpha(B1.1.177)). An interaction term between viral load and lateral flow device is included to allow the shape of the curves plotted to vary by device.

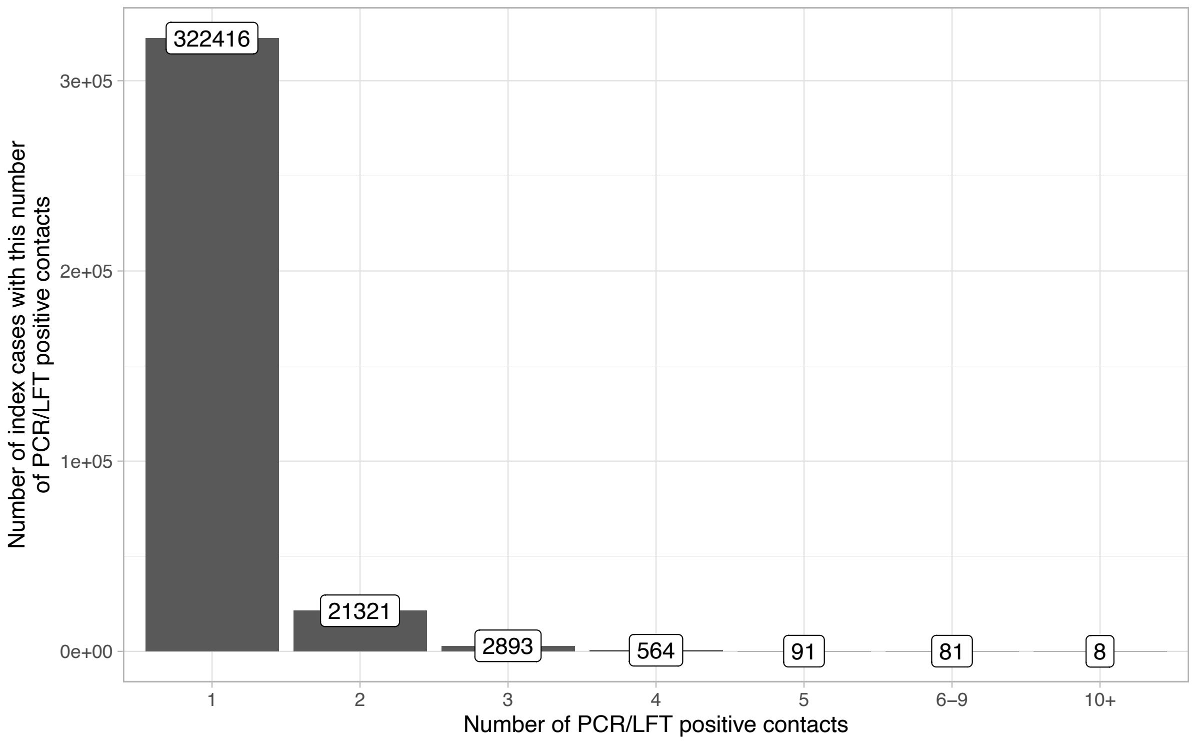


**Figure S3. Number of PCR/LFD-positive contacts per index case in national contact testing data**.

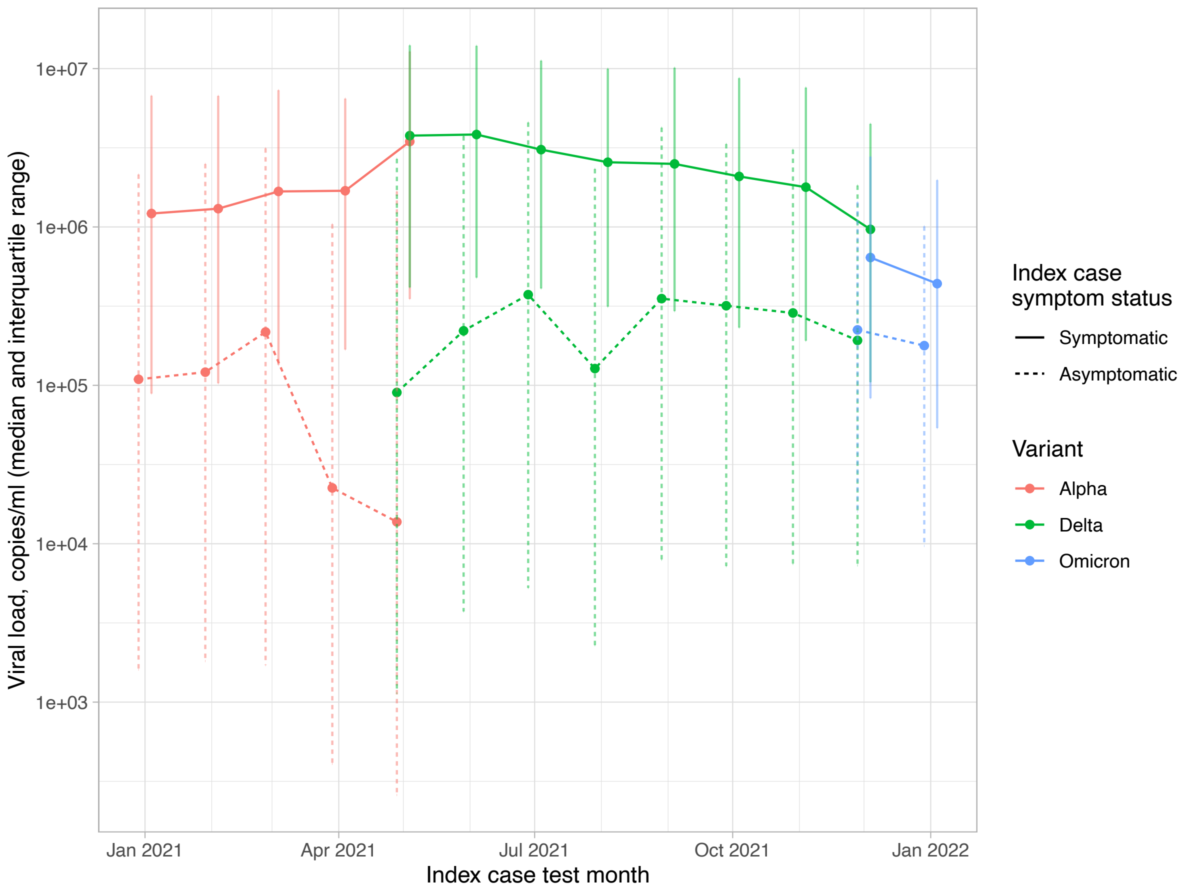


**Figure S4. Index case viral load in probable case-contact transmission pairs.** The median viral load is plotted with “error bars” indicating the interquartile range. Data are plotted aggregated by month with lines coloured by estimated variant, and the line type indicated index case symptom status

### Supplementary Tables

| **Lab** | **Ct (denoted x) to $\log_{10}$(viral load, copies per ml, denoted y)** | | | | |
|---|---|---|---|---|---|
| | **ORF1ab** | **N-Gene** | **S-Gene** | **E-Gene** | **RdRp** |
| **Randox Assay AQP1-A** | y = -0.3065x + 12.477 | - | - | y = -0.3103x + 12.850 | - |
| **Randox Assay PE** | y = -0.2909x + 11.921 | y = -0.3355x + 13.334 | - | - | - |
| **Alderley Park** | y = -0.3035x + 11.599 | y = -0.3120x + 11.881 | - | - | - |
| **Glasgow** | y = -0.3050x + 11.372 | y = -0.3096x + 11.449 | y = -0.2894x + 11.221 | - | - |
| **Milton Keynes** | y = -0.3181x + 11.859 | y = -0.3241x + 12.119 | y = -0.3641x +13.372 | - | - |
| **HSL UCL** | - | y = -0.2915x + 14.049 | - | - | - |
| **Newcastle** | y= -0.2831x + 11.354 | y = -0.3161x + 11.882 | y = -0.3857x + 14.003 | - | - |
| **Plymouth** | y = -0.2971x + 12.649 | y = -0.3441x + 14.637 | - | - | - |

**Table S1. Conversion formulae from SARS-CoV-2 PCR results to viral load**. Results were determined using the Qnostics SCV2AQP01 quantitative SARS-CoV-2 standards panel, and linear regression models. For LFD performance assessments the mean viral load was calculated across all detected targets. For transmission analyses, index case viral loads were calculated using the mean viral load based on Ct values for the ORF1ab and N genes.

| Scenario | Total population | Population prevalence | Prevalence of symptoms due to other reasons | Number symptomatic for any reason | Proportion of infections symptomatic | Proportion of symptomatic cases who transmit | Proportion of asymptomatic cases who transmit | Number of potential symptomatic transmitters | Number of potential asymptomatic transmitters | LFD sensitivity in symptomatic transmitters | LFD sensitivity in asymptomatic transmitters | Number needed to test to detect one symptomatic transmitter | Number needed to test to detect one asymptomatic transmitter |
|---|---|---|---|---|---|---|---|---|---|---|---|---|---|
| Baseline | 100000 | 1% | 0.5% | 1000 | 50% | 6% | 4% | 30 | 20 | 79% | 57% | 42 | 8684 |
| Increased prevalence | 100000 | **2%** | 0.5% | 1500 | 50% | 6% | 4% | 60 | 40 | 79% | 57% | 32 | 4320 |
| | 100000 | **5%** | 0.5% | 3000 | 50% | 6% | 4% | 150 | 100 | 79% | 57% | 25 | 1702 |
| Increased symptoms for other reasons | 100000 | 1% | **1.0%** | 1500 | 50% | 6% | 4% | 30 | 20 | 79% | 57% | 63 | 8640 |
| | 100000 | 1% | **5.0%** | 5500 | 50% | 6% | 4% | 30 | 20 | 79% | 57% | 232 | 8289 |
| Increased transmission | 100000 | 1% | 0.5% | 1000 | 50% | **20%** | **10%** | 100 | 50 | 79% | 57% | 13 | 3474 |
| Comparable LFD performance regardless of symptoms | 100000 | 1% | 0.5% | 1000 | 50% | 6% | 4% | 30 | 20 | 79% | 79% | 42 | 6266 |
| Perfect LFD performance | 100000 | 1% | 0.5% | 1000 | 50% | 6% | 4% | 30 | 20 | **100%** | **100%** | 33 | 4950 |
| | 100000 | **5%** | 0.5% | 3000 | 50% | 6% | 4% | 150 | 100 | **100%** | **100%** | 20 | 970 |

**Table S2. Number of LFD tests needed to detect symptomatic and asymptomatic cases that would otherwise go on to transmit**. Different scenarios are shown for an example population of 100,000 people, the factors changed in each scenario are shown in bold. For illustrative purposes, a total of 50% of infections are assumed to be asymptomatic. We assume that 6% of index cases went on to transmit and that asymptomatic cases were around 0.7-times as infectious as symptomatic cases, based on estimates from this study, accepting these rates depend on tests being sought by contacts between 1 and 10 days following the index cases' diagnosis and are therefore likely to be somewhat underestimated (a scenario with higher transmission rates is also shown). Estimates of LFD sensitivity in symptomatic and asymptomatic transmitters are taken from this study.

### References

1. Eyre DW, Taylor D, Purver M, Chapman D, Fowler T, Pouwels KB, et al. Effect of Covid-19 Vaccination on Transmission of Alpha and Delta Variants. New Engl J Med. 2022;386: 744–756. doi:10.1056/nejmoa2116597

2. Lee LYW, Rozmanowski S, Pang M, Charlett A, Anderson C, Hughes GJ, et al. SARS-CoV-2 infectivity by viral load, S gene variants and demographic factors and the utility of lateral flow devices to prevent transmission. Clin Infect Dis. 2021; ciab421-. doi:10.1093/cid/ciab421
